# Continuous monitoring of SARS-CoV-2 RNA in urban wastewater from Porto, Portugal: sampling and analysis protocols

**DOI:** 10.1101/2021.04.06.21254994

**Authors:** Maria Paola Tomasino, Miguel Semedo, Pedro Vieira, Elza Ferraz, Adelaide Rocha, Maria F. Carvalho, Catarina Magalhães, Ana P. Mucha

**Affiliations:** Interdisciplinary Centre of Marine and Environmental Research (CIIMAR/CIMAR), University of Porto, Matosinhos, Portugal; AEdPorto (Empresa de Águas e Energia do Município do Porto, EM), Porto, Portugal; Institute of Biomedical Sciences Abel Salazar, University of Porto, Porto, Portugal; Faculty of Sciences, University of Porto, Porto, Portugal

## Abstract

Research on the emerging COVID-19 pandemic is demonstrating that wastewater infrastructures can be used as public health observatories of virus circulation in human communities. Important efforts are being organized worldwide to implement sewage-based surveillance of SARS-CoV-2 that can be used for preventive or early warning purposes, informing preparedness and response measures. However, its successful implementation requires important and iterative methodological improvements, as well as the establishment of standardized methods. The aim of this study was to develop a continuous monitoring protocol for SARS-CoV-2 in wastewater, that could be used to model virus circulation within the communities, complementing the current clinical surveillance. Specific objectives included (1) optimization and validation of a sensitive method for virus quantification; (2) monitoring the time-evolution of SARS-CoV-2 in wastewater from two wastewater treatment plants (WWTPs) in the city of Porto, Portugal. Untreated wastewater samples were collected weekly from the two WWTPs between May 2020 and March 2021, encompassing two COVID-19 incidence peaks in the region (mid-November 2020 and mid-January 2021). In the first stage of this study, we compared, optimized and selected a sampling and analysis protocol that included RNA virus concentration through centrifugation, RNA extraction from both liquid and solid fractions and quantification by reverse transcription quantitative PCR (RT-qPCR). In the second stage, we used the selected methodology to track SARS-CoV-2 in the collected wastewater over time. SARS-CoV-2 RNA was detected in 39 and 37 out of 48 liquid and solid fraction samples of untreated wastewater, respectively. The copy numbers varied throughout the study between 0 and 0.15 copies/ng RNA and a good fit was observed between the SARS-CoV-2 RNA concentration in the untreated wastewater and the COVID-19 temporal trends in the study region. In agreement with the recent literature, the results from this study support the use of wastewater-based surveillance to complement clinical testing and evaluate temporal and spatial trends of the current pandemic.

## 1. Introduction

Municipal WWTPs have an important function in modern urban life, by promoting the degradation of organic waste, removal of phosphorus and nitrogen, and the reduction of pathogens before the release of treated water into the surrounding environment. Thus, these systems became crucial to maintain public health in urban environments and to reduce the impact of dense populated areas in the natural ecosystems. However, because WWTPs receive waste effluents from different uses (e.g. hospitals, industry, agricultural and urban), they are also considered major hotspots of nutrient enriched waters and of multiple biological and chemical pollutants. Among biological contaminants, microbial pathogens are a major problem associated with wastewaters, concerning ecology and human health risks (Bogler et al., 2020). This underscores a challenge regarding improvement on the disinfection efficiency of WWTPs effluents and of the resulting residues that will be reused (Bogler et al., 2020). On the other hand, the fact that sewage functions as a depository of human excretions, can make these systems a promising observatory of the incidence of pathogens in the population. Thus, it has been argued that wastewaters contain valuable information that can be useful to forecast community health risk (Venkatesan and Halden, 2014).

The idea that wastewater can be a valuable epidemiological data source was particularly reinforced during the current COVID-19 pandemic, where several studies demonstrated the presence of SARS-CoV-2 in wastewater (Ahmed et al., 2020a; Gonzalez et al., 2020; Kumar et al., 2020; Medema et al., 2020; Peccia et al., 2020; Randazzo et al., 2020). In fact, some recent studies found a clear positive relation between virus concentration in wastewater and the reported COVID-19 cases in the community (Medema et al., 2020; Peccia et al., 2020). Thus, the relevance of monitoring SARS-CoV-2 concentrations in wastewater to track COVID-19 became a priority for infection surveillance at the population level (Peccia et al., 2020). As recently recommended by the European Commission, “*Surveillance of SARS-CoV-2 in wastewater can provide important complementary and independent information to the public health decision-making process in the context of the ongoing COVID-19 pandemic. As a consequence, wastewater monitoring needs to be included more systematically in the national testing strategies for the detection of the SARS-CoV-2 virus*.” (Commission, 2021). Notwithstanding, handling and processing a complex matrix like sewage samples is highly challenging and the implementation of a successful SARS-CoV-2 monitoring plan requires iterative methodological improvements, as well as the establishment of standardized methods, to support the assessment of geographic and temporal trends.

In the present study, we describe the optimization steps for a monitoring program to detect and quantify SARS-CoV-2 RNA in untreated wastewater, and report, on a weekly basis, the viral loads between September 2020 to March 2021 in the two WWTPs of the city of Porto, Portugal. By including different sampling, RNA isolation, and SARS-CoV-2 detection methodologies, this study provides relevant information that can be used in future monitoring programs for sewage-based surveillance of SARS-CoV-2.

Wastewater treatment in the city of Porto is carried out by two WWTPs: Sobreiras and Freixo (Figure S1). The Sobreiras WWTP, which serves the westernmost part of the city (including the largest hospitals), was designed to treat an average daily flow of 54,000 m^3^ of wastewater and to serve an equivalent population of 200,000 inhabitants. In turn, the Freixo WWTP treats wastewater produced in the eastern part of the city of Porto and also part of the wastewater generated in the municipality of Gondomar. This facility has the capacity to receive and treat 34,900 m^3^ daily and to serve 170,000 equivalent inhabitants. Currently, and more specifically in the period under study, an average of 33,000 m^3^ and 24,000 m^3^ of wastewater per day flowed and were treated, respectively, in the Sobreiras and Freixo WWTPs. In terms of organic matter, it should be noted that although the concentrations are similar, the affluent loads are higher in the Sobreiras WWTP (higher average daily flow).

During the period of this study, the number of new COVID-19 cases per day in the city of Porto varied between 0 - 455 and two incidence peaks were observed, one in mid-November 2020 and another in mid-January 2021. To our knowledge, this is the first published report on SARS-CoV-2 monitoring in Portuguese WWTPs, providing a novel data source that will contribute to the global effort of monitoring SARS-CoV-2 circulation in human communities.

## 2. Material and Methods

Composed 24h raw sewage samples were weekly collected from two WWTPs (Sobreiras and Freixo) in the metropolitan area of Porto (Portugal) during 44 weeks, from May 2020 to March 2021, with a total of 81 samples processed. Over this period, the detection of SARS-CoV-2 in sewage followed two stages (**Figure 1**). The first one (May - September 2020) included the protocol optimization by testing different procedures of sample precipitation and filtration, RNA extraction and virus detection. During the second stage, from September 2020 to March 2021, the most successful protocol was selected and used to weekly monitor SARS-CoV-2 in both liquid (supernatant) and solid (suspended particles) fractions of untreated wastewater. Although in the present study we report results until the beginning of March 2021, this second stage is still ongoing and it is currently providing a time-evolution monitoring of SARS-CoV-2 in Sobreiras and Freixo WWTPs.

**Figure 1.**
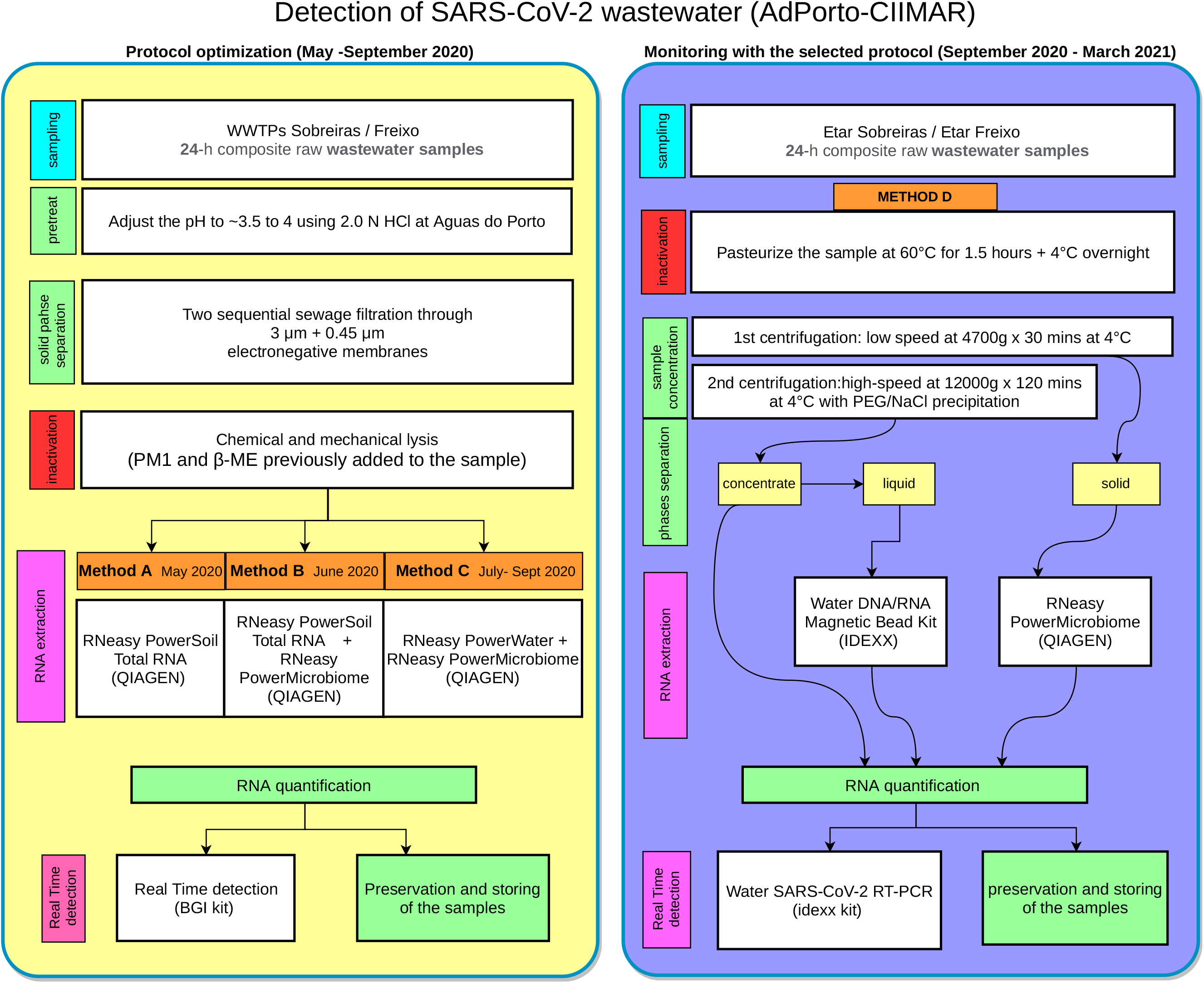
Workflow of SARS-CoV-2 wastewater detection (AdPorto-CIIMAR): on the left, the first stage of protocol optimization (May - September 2020); on the right, the second stage of SARS-CoV-2 weekly monitoring in both liquid and solid fractions (September 2020 to March 2021) in Sobreiras and Freixo WWTPs.

### Protocol optimization (May - September 2020)

#### Wastewater sampling and pre-treatment

Sewage samples were collected during 20 weeks from Sobreiras and Freixo WWTPs (with the exception of the first six weeks when only Sobreiras was sampled). Pre-treatment of the samples included the adjustment of their pH to ∼3.5/4 using 2.0 N HCl. Pre-treatment acidification, as well as liquid phase separation and RNA extraction were performed according to (Ahmed et al., 2015, 2020a).

#### Liquid phase separation

A variable volume of acidified sewage sample (ranging from 10 to 80 ml) was sequentially filtered through 3 μm + 0.45 μm pore-size, 90 mm diameter electronegative membranes (SSWP04700 and HAWP04700; Merck Millipore), just after sample collection at WWTPs laboratories.

#### Virus inactivation

Immediately after filtration, electronegative membranes were accommodated in the respective collection tubes containing already the solutions for the inactivation and chemical lysis of the virus, namely PM1 (RNeasy PowerMicrobiome Kit Component - Qiagen, GMBH, Germany) and β-mercaptoethanol (Sigma). Samples were then transported to CIIMAR laboratories, in ice chests, for further molecular analysis and storage.

#### RNA extraction

RNA was extracted directly from the electronegative membranes using three different procedures. *Method A_May 2020*: RNeasy Power Soil Kit (Qiagen) was used according to the manufacturer’s protocol; *Method B_ June 2020*: 15-mL falcon tubes from RNeasy PowerSoil Kit (Qiagen) were used to accommodate the electronegative membranes. Then RNA was extracted following RNeasy PowerMicrobiome Kit; *Method C_July-September 2020*: 5-mL bead tubes were used to accommodate the electronegative membranes following steps 1-6 of RNeasy PowerWater Kit. From here, RNA was extracted using RNeasy PowerMicrobiome Kit. Elution of RNA was done in a 100 μl elution buffer for all RNA extraction methods.

#### RT-qPCR virus detection and RNA quantification and preservation

The total RNA extracted was quantified by Nanodrop and divided in two aliquots. One aliquot was reverse transcribed to cDNA (using QuantiNova Reverse Transcription Kit, Quiagen) and stored at − 80°C. The other aliquot was processed through RT-qPCR by using BGI’s Real-Time Fluorescent RT-PCR kit for detecting 2019-nCoV (SARS-CoV-2) (IVD 127 & CE marked; Catalogue No. MFG030010). Each sample was analysed in duplicate wells in a StepOnePlus(tm) Real-Time PCR System. Both positive-control (all RT-qPCT reagents plus SARS-CoV-2 rRNA) and negative-control (all RT-qPCR reagents without template) assays were included for quality control.

### Continuous Monitoring with the Selected Protocol (September 2020 - March 2021)

#### Wastewater sampling

Sewage sampling (24 h composed samples) was weekly performed from Sobreiras and Freixo WWTPs between September 23th (week 20) to March 10th (week 44). Samples were then directly transferred to CIIMAR laboratories under cool conditions and processed on the same day of sampling (Method D; Figure 1).

#### Inactivation

A total volume of 500 ml composed sewage samples was pasteurized at 60 °C for 90 min to inactivate SARS-CoV-2, in order to increase the safety of the laboratory personnel during sample handling (La Rosa et al., 2020; F. Wu et al., 2020). Samples were kept overnight at 4 °C until further processing that started the next morning.

#### Phase separation, concentration, RNA extraction

A volume of 105 ml of pasteurized sewage was divided in three 50 ml falcon tubes (containing 35 ml of sewage each) and centrifuged at 4,700 g for 30 min at 4°C. The resulting supernatant (*liquid*) and the pellet (*solid*) fractions were carefully separated. RNA was directly extracted from the solid phase (pellet) using RNeasy PowerMicrobiome Kit (Qiagen). The supernatant was transferred to new falcon tubes for PEG/NaCl precipitation to concentrate the viruses from aqueous matrix (Ikner et al., 2012; Wyn-Jones and Sellwood, 2001), which was carried out by adding 3.8 g of Polyethylene glycol 8000 (8% w/v, Millipore Sigma) and 0.8 g of NaCl (0.3 M, Millipore Sigma) to the supernatant. Samples were then centrifuged at high speed (12000 g, 2 hours at 4°C) and the resulting pellet was resuspended in 400 μl of RNA free water, obtaining what we call the ‘*concentrate’* phase. The RNA was extracted from 200 μl of this ‘*concentrate’* phase by using Water DNA/RNA Magnetic Bead Kit (IDEXX Laboratories, Inc., Westbrook, ME) (Pecson et al., 2021). RNA was eluted in 100 µl elution buffer in both solid and liquid wastewater fractions.

#### RT-qPCR virus detection and RNA quantification and preservation

The total RNA extracted from the three matrices (concentrate, liquid, solid) was quantified by Nanodrop and divided in two aliquots. One aliquot was reverse transcribed to cDNA (using QuantiNovaTM Reverse Transcription Kit, Qiagen) and stored at −80°C. The other aliquot was processed by using the Water SARS-CoV-2 RT-PCR ready-to-use kit (IDEXX Laboratories, Inc., Westbrook, ME), designed to target both the 2019-nCoV_N1 and 2019-nCoV_N2 markers of the virus. Each sample was quantified by RT-qPCR in duplicate wells in a Step One Plus real-time PCR system. A positive detection was considered when the Ct’s of both replicates from the same sample were < 42. Both positive-control and negative-control assays were performed for quality control as previously described. RT-qPCR standards were prepared through a serial dilution of head-inactivated SARS-CoV-2 (ATCC number: VR-1986HK(tm)) carrying the target genes. Two standard curves were prepared in two independent qPCR runs and the parameters Y-intercept and slope were averaged and used to estimate virus copy numbers for all weekly measurements. Gene copy number per PCR well was calculated from the standard curve according to the equation (1):

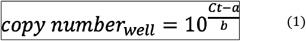

Where Ct corresponds to the threshold cycle of the sample, and *a* and *b* correspond to the Y-intercept and slope of the logarithmic standard curve, respectively. Copy numbers per well were then converted to copy numbers per ng of RNA by dividing the total RNA present in the 5 μL of each sample added to the well.

### Statistical analysis and data visualization

Differences in total RNA concentration, A260/A230, A260/A280, and the Ct value between the liquid and solid phases at each WWTP were compared with a two-sample Mann-Whitney-Wilcoxon test (non-parametric). Simple linear regressions were used to assess the relationship between Ct values and total RNA concentrations as well as between the SARS-CoV-2 copy numbers/ng RNA and the COVID-19 moving average at each sampling day. Significant relationships for all tests were considered at ? < 0.01. All statistical analyses and plots were conducted in the R environment (version 3.2.2. Copyright 2015 The R Foundation for Statistical Computing), using base R and the “ggplot2” package.

## 3. Results and Discussion

### Comparison between methods

The concentration of RNA extracted from the collected wastewater samples using the different methods, as well as the respective qPCR Ct values obtained during the first stage of this study (protocols optimization) are shown in **Table 1**. In the first sampling week (2020-05-14), we compared method A (PowerSoil) with method B (PowerSoil + PowerMicrobiome). Method B recovered a higher concentration of total RNA (55.0 ng/µL) than method A (35.7 ng/µL), but both methods had negative results in the qPCR for this period of sampling. In the third week (2020-05-28), we included method C (PowerWater + PowerMicrobiome), previously used to detect SARS-CoV-2 RNA in untreated wastewater (Ahmed et al., 2020a). Despite the qPCR detection remaining negative, method C doubled the amount of total RNA isolated (161.0 ± 13.8 ng/µL), when compared to method B (81.9 ± 41.5 ng/µL) (Table 1). It is reasonable to expect that higher total RNA concentrations would increase chances of viral RNA detection in downstream qPCR, so method C was selected for the following weeks. To our knowledge, the recent literature concerning viral RNA detection in wastewater do not include information about total RNA yields of the different extraction protocols (e.g. (Ahmed et al., 2020a, 2020b; Gonzalez et al., 2020; Kumar et al., 2020; Medema et al., 2020; Peccia et al., 2020; Randazzo et al., 2020; Sherchan et al., 2020; F. Wu et al., 2020)).

**Table 1.**
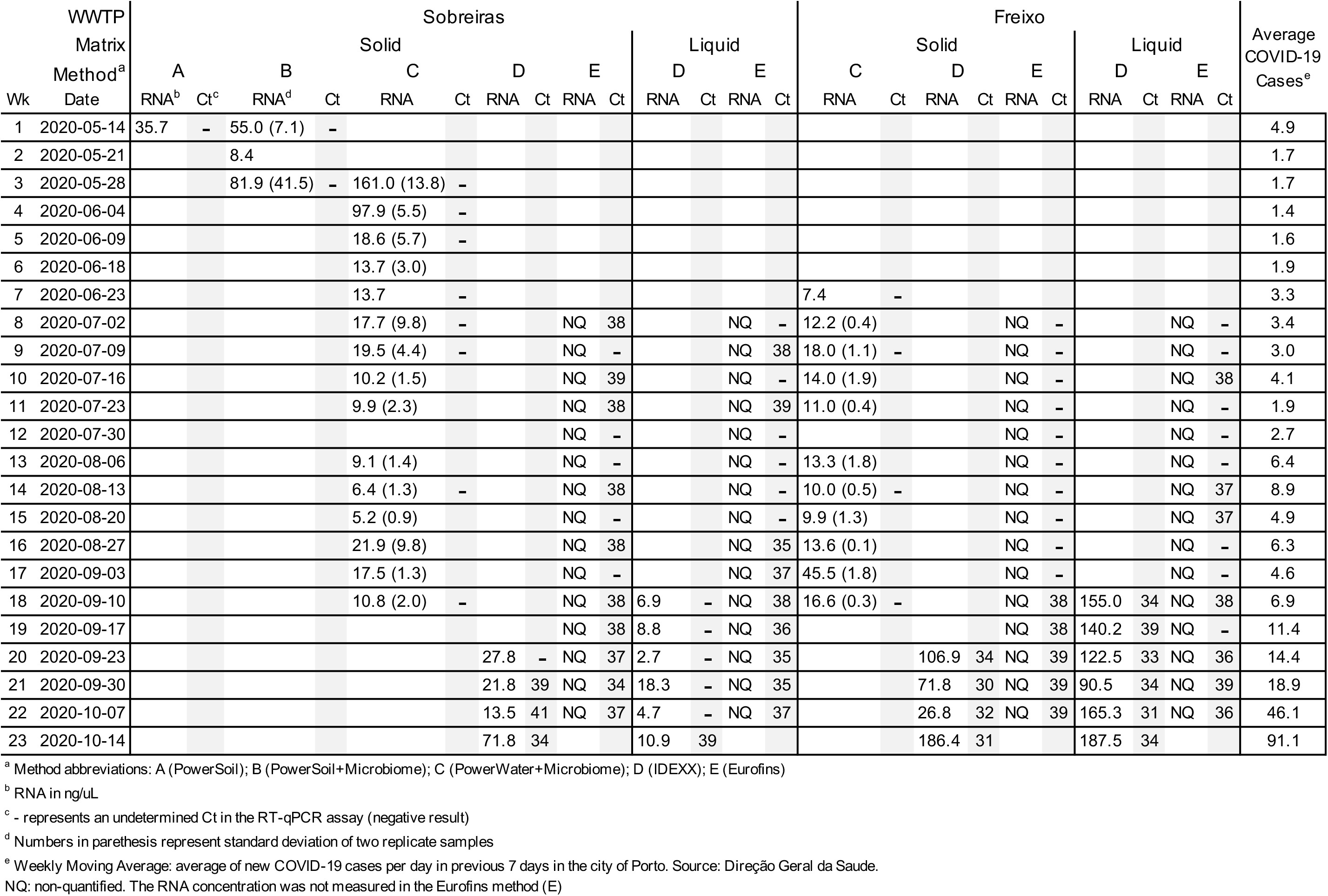
Extracted total RNA concentrations and qPCR Ct values of the different SARS-CoV-2 RNA detection methods tested during the first stage of this study (protocol optimization).

From week 3 (2020-05-28) to week 17 (2020-09-03), we used method C due to its high RNA recovery and its previous validation for sewage samples (Ahmed et al., 2020a). However, we did not detect the presence of SARS-CoV-2 RNA in any samples analyzed during this period. In parallel to our analysis, samples were sent to an external service provider (Eurofins) that was able to detect a total of 12 positive results in the same period (30% of tested samples) (*method E* - **Table 1**). However, it is important to note that the obtained Ct values were not consistent and were close to the qPCR threshold reported for the Eurofins method (Ct = 38; (Jørgensen et al., 2020)) and to the threshold recommended by the European Commission (Ct = 40) to report a sample as positive (Commission, 2021). This inconsistency is probably due to the relatively low number of COVID-19 cases in the city of Porto before mid-October 2020. Since most positive results with the Eurofins method were detected in the liquid phase (7 out of 12 positives) and only this phase had positive detection in both WWTPs (Freixo and Sobreiras), we tested an additional method for detection of the virus in the liquid phase (*method D* - IDEXX + PowerMicrobiome; **Figure 1**). Here we included a step of high-speed centrifugation with viral RNA precipitation/concentration that have been shown to successfully detect SARS-CoV-2 RNA in wastewater samples during the COVID-19 pandemic (Ahmed et al., 2020b; Kumar et al., 2020; F. Wu et al., 2020; Zhang et al., 2020).

On week 18 (2020/09/10), we detected SARS-CoV-2 RNA in one of the tested samples (Freixo WWTP, liquid phase) with method D, with a Ct of 34 and a high RNA extraction yield. On the same sampling day, method C gave negative results for both WWTPs. Since then, we started using only method D, that coupled the IDEXX magnetic bead kit to recover RNA in the liquid phase with the PowerMicrobiome kit to isolate RNA from the solid phase (Figure 1). From week 21 (2020/09/30) onwards, we started detecting SARS-CoV-2 RNA in both WWTPs and on week 23 (2020/10/14) we detected SARS-Cov-2 RNA for the first time in all samples collected for this study. The improved detection in mid-October was concomitant with an increase in the number of COVID-19 cases in the region (**Table 1**). From then on, we used only method D to track the SARS-CoV-2 in the liquid and solid phases of wastewater from Sobreiras and Freixo WWTPs on a weekly basis.

It is also important to mention that, besides the solid and liquid fractions, we occasionally analyzed the raw centrifugate obtained after the concentration step, that we called ‘*concentrate’*. We tested this *concentrate* recovered from both WWTPs directly in RT-qPCR, skipping the step of RNA extraction. Although we registered positive detection of SARS-CoV-2 RNA in this matrix, we found that RNA yield and Ct values were not consistent between replicates compared with the ones where the RNA extraction step was included. Most probably, impurities of this concentrate limited the efficiency of RT-qPCT step (data not shown).

### SARS-CoV-2 detection in liquid and solid wastewater fractions

Different procedures are reported in the literature to detect SARS-CoV-2 in both liquid and solid fractions of the wastewater matrix (Michael-Kordatou et al., 2020). However, there is still a lack of consensus in literature about which matrix shows better performance for SARS-CoV-2 detection (Peccia et al., 2020; F. Wu et al., 2020). In the second stage of this study, we analyzed the liquid and solid phases of 48 untreated wastewater samples from the two WWTPs over 24 weeks (from September 23rd, 2020, to March 10th, 2021), by applying Method D (Figure 1, Table S1). Each sample was quantified by RT-qPCR in duplicate wells, in a total of 192 tests (96 on liquid and 96 on solid phases). Overall, during this period, SARS-CoV-2 was detected in 39 out of 48 samples (81%) of liquid fractions and 37 out of 48 samples (77%) of solid fractions from the untreated wastewater. Besides testing the untreated wastewater, we also quantified SARS-CoV-2 RNA in 8 samples of treated wastewater (four sampling weeks from both WWTP), which gave negative results for both plants (data not shown). This is in contrast with some recent studies that reported the detection of SARS-CoV-2 RNA by RT-qPCR in treated wastewaters and even in rivers (Nasseri et al., 2021; Rimoldi et al., 2020; Wurtzer et al., 2021, 2020), although near the quantification limits. These findings could be important to indicate if the wastewater treatment technologies are being efficient in the removal of SARS-CoV-2 and could provide a comprehensive view on the fate of SARS-CoV-2 along the entire chain of WWTPs.

In general, the solid phase showed higher RNA recovery (although with a greater variation) compared with the liquid fraction in both WWTPs. In Freixo, total RNA yield average was 95.1 ± 33 ng/µl in the liquid and 194 ± 173 ng/µl in the solid phase (n= 48; p= 0.08), while in Sobreiras RNA yield was 16 ± 7 ng/µl in liquid and 200 ± 255 ng/µl in solid phases (n= 48; p= 0.004) (**Figure 2A**). RNA quality was also evaluated to assess phenol/carbohydrates contamination (260/230) and protein contamination (260/280). Liquid phase samples presented a lower 260/230 ratio than solid phase samples in both the WWTPs. In Freixo, 260/230 average was 0.6 ± 0.2 in liquid and 2 ± 0.5 in solid samples (n= 48; p= 0.00003), while in Sobreiras was 0.8 ± 0.5 in liquid and 1.9 ± 0.6 in solid samples (n= 48; p=0.00002) (**Figure 2B**). Similar trends were registered for RNA A260/280 absorbance ratio, with values >1.9 in the solid matrices of both WWTPs, showing lower protein contamination than liquid matrices. In Freixo, 260/280 average was 1.5 ± 0.1 in liquid and 2.2 ± 1.1 in solid samples (n= 48; p=0.0001), while in Sobreiras was 1.4 ± 0.2 in liquid and 2 ± 0.3 in solid samples (n=48; p=0.0000005, **Figure 2C**). These findings show that RNA extracted from the liquid matrix could contain higher residual contaminants (protein, carbohydrate, residual phenol or salts), affecting RNA quality with a possible influence on the accuracy and reliability of downstream applications. Despite the differences in the quantity and quality of the RNA between liquid and solid phases, the Ct values overlapped between the liquid and solid matrices (in Freixo n=96; p=0.57; in Sobreiras n=96; p=0.2) with an average of 33.7 ± 3.7 for the liquid samples and 34.2 ± 5 for the solid samples (**Figure 2D**). These results suggest that, in our study, virus detection does not seem to be solely affected by nucleic acid extraction yield and purity of RNA. To confirm this indication, we tested the linear relationship between total RNA extracted and the RT-qPCR Ct value (**Figure 3**). Results showed no significant relationship between Ct values and RNA concentrations in both phases for neither Freixo nor Sobreiras WWTP. In fact, Ct values below 29 could be observed with total RNA concentrations as low as 11.7 ng/µL. The lack of a significant relationship between Ct and RNA concentrations observed in our study underscores that, for this range of RNA concentrations, the extracted amounts are not critical for the Ct value observed in the qPCR assay.

**Figure 2.**
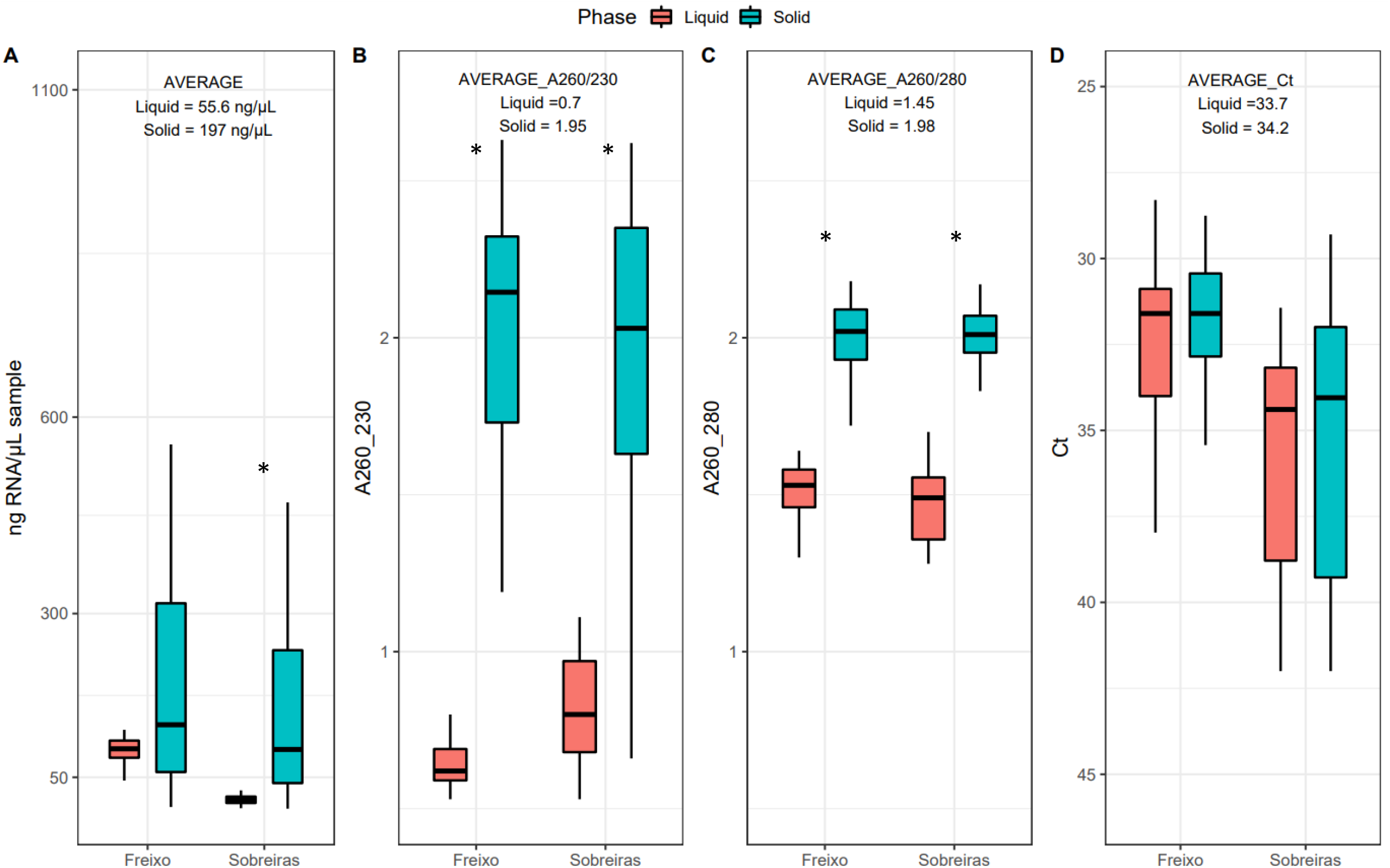
Box-plots representing the total RNA yield (A), RNA purity by A260/230 ratio (B), by A260/280 ratio (C), and the qPCR Ct values (D) in liquid and solid fractions of Sobreiras and Freixo WWTPs. The boxes represent the interquartile range (difference between the upper 75% and lower quartile 25%); the inner box lines represent the medians; *p<0.01 (Mann-Whitney-Wilcoxon test).

**Figure 3.**
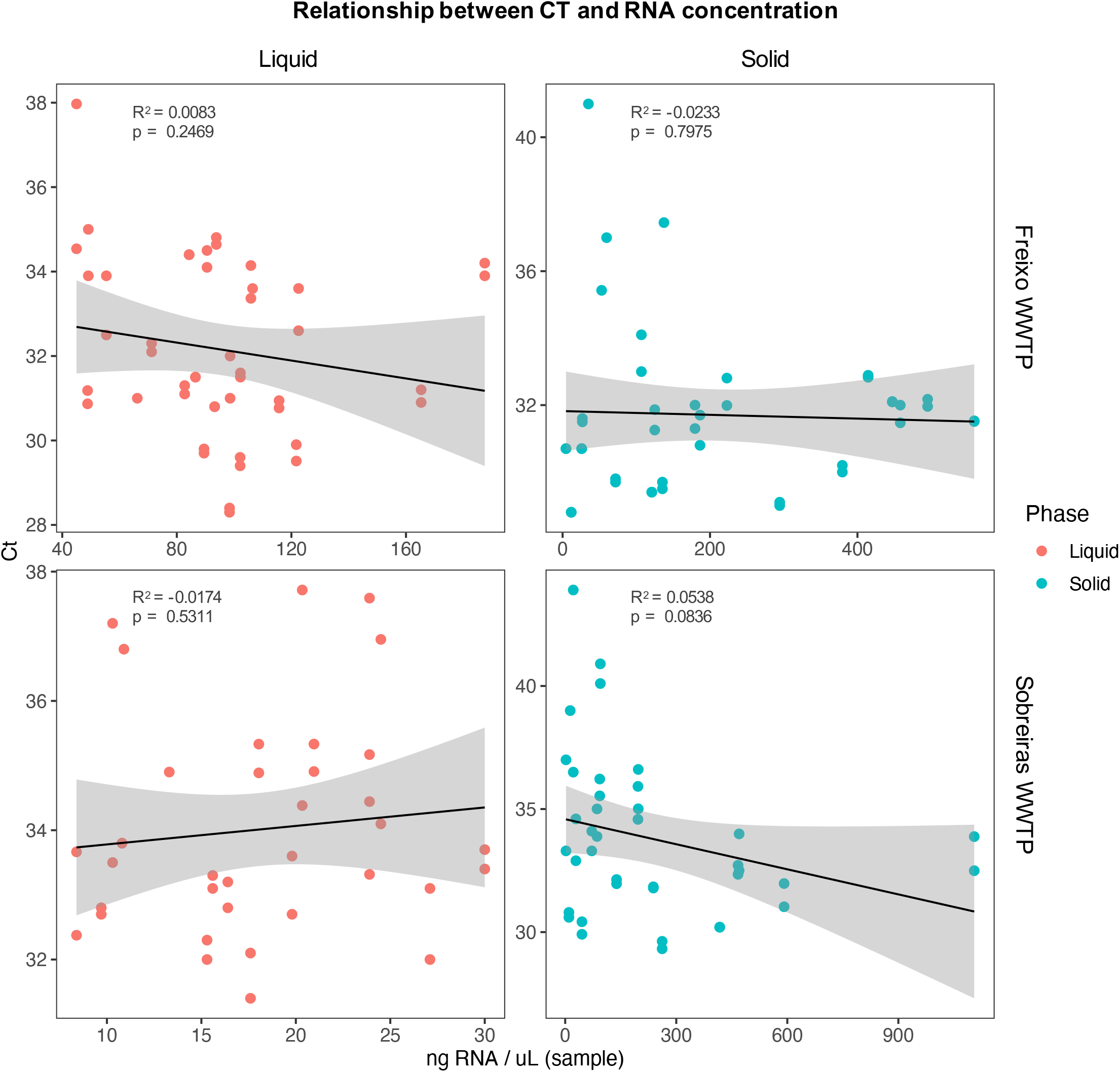
Linear regressions between SARS-CoV-2 qPCR Ct values and total RNA concentrations extracted from the liquid and solid phases of untreated wastewater from Freixo and Sobreiras WWTPs. The shaded area represents the 95% confidence interval of the linear regression predictions.

Furthermore it is important to highlight the heterogeneity of SARS-CoV-2 detection in the two different WWTPs that can represent a limiting factor for sewage virus monitoring. In our results, we observed that considering the liquid phase, SARS-CoV-2 was detected in 91% of samples (22 out of 24) from Freixo and 70% of samples (17 out of 24 samples) from Sobreiras; while considering the solid phase, 79% of samples from Freixo (19 out of 24) and 75% (18 out of 24) from Sobreiras were positive for the virus. Differences observed between the two WWTPs could depend on the different daily flow of the two WWTPs and on the different physicochemical conditions of the wastewaters (i.e pH, temperature, fraction of particulate matter, presence of micropollutants, etc.) that may have a significant impact on the stability and survival of the virus (Auffret et al., 2019; Michael-Kordatou et al., 2020). However, these results could also reflect the difference of SARS-CoV-2 dynamics in the populations serving the two WWTPs. For this reason, further investigation would be useful to compare the viral load to the number of infected persons in the catchment area of each WWTP, but this is out of the scope of this work. Notwithstanding the WWTP heterogeneity, our data show the feasibility of measuring SARS-CoV-2 in both suspended solids and liquid fractions of wastewater. While detection in liquid phase showed less variability in both WWTPs (**Figure 2**), probably due to its more homogeneous nature, we believe that a parallel detection on solid and liquid fractions should be carried out to improve recovery efficiency of enveloped viruses from raw wastewater (La Rosa et al., 2020; F. Wu et al., 2020; Ye et al., 2016).

### Time-evolution of SARS-CoV-2 in wastewater

In order to evaluate the future applicability of the developed monitoring protocol for the establishment of quantitative predictions of SARS-CoV-2 occurrence in the population, it is important to compare the results obtained over time with the epidemiological context of COVID-19 in the region. Two epidemic peaks were registered in the city of Porto during this study, one in mid-November 2020 and the other one in mid-January 2021 (**Figure 4**). The abundance of SARS-CoV-2 RNA in untreated wastewater between late September 2020 and mid-March 2021 is displayed in **Figure 5**. In general, we observed a good fit between the SARS-CoV-2 gene copy numbers in the analyzed wastewater and the COVID-19 incidence peaks in the region, especially with the peak observed in mid-November. The highest SARS-CoV-2 copy numbers observed for both WWTPs in the liquid and solid phases were coincident with the mid-November peak, with the exception of a few higher values observed during the mid-January peak in the liquid phase of Sobreiras WWTP. Studies performed during the current pandemic in different world regions have also shown a good fit between COVID-19 cases and the SARS-CoV-2 detection in wastewater (Ahmed et al., 2020a; Gonzalez et al., 2020; Haramoto et al., 2020; Kumar et al., 2020; Medema et al., 2020; Peccia et al., 2020; Sherchan et al., 2020), which is expected due to the reported presence of SARS-CoV-2 RNA in fecal samples from COVID-19 patients (Y. Wu et al., 2020). Other studies have found even higher SARS-CoV-2 RNA numbers in wastewater than expected based on clinically-confirmed cases (F. Wu et al., 2020). The apparently weaker fit between new COVID-19 cases and the copy numbers in the wastewater during the second peak (especially in the solid phase) may be due to different factors, such as a dilution effect associated with precipitation, the effect of the lockdown on the ration between resident/nonresident population discharging to the city sewage system, and/or different clinical testing magnitude. Despite the reasons behind these differences being beyond the scope of this communication, they can be investigated in future studies.

**Figure 4.**
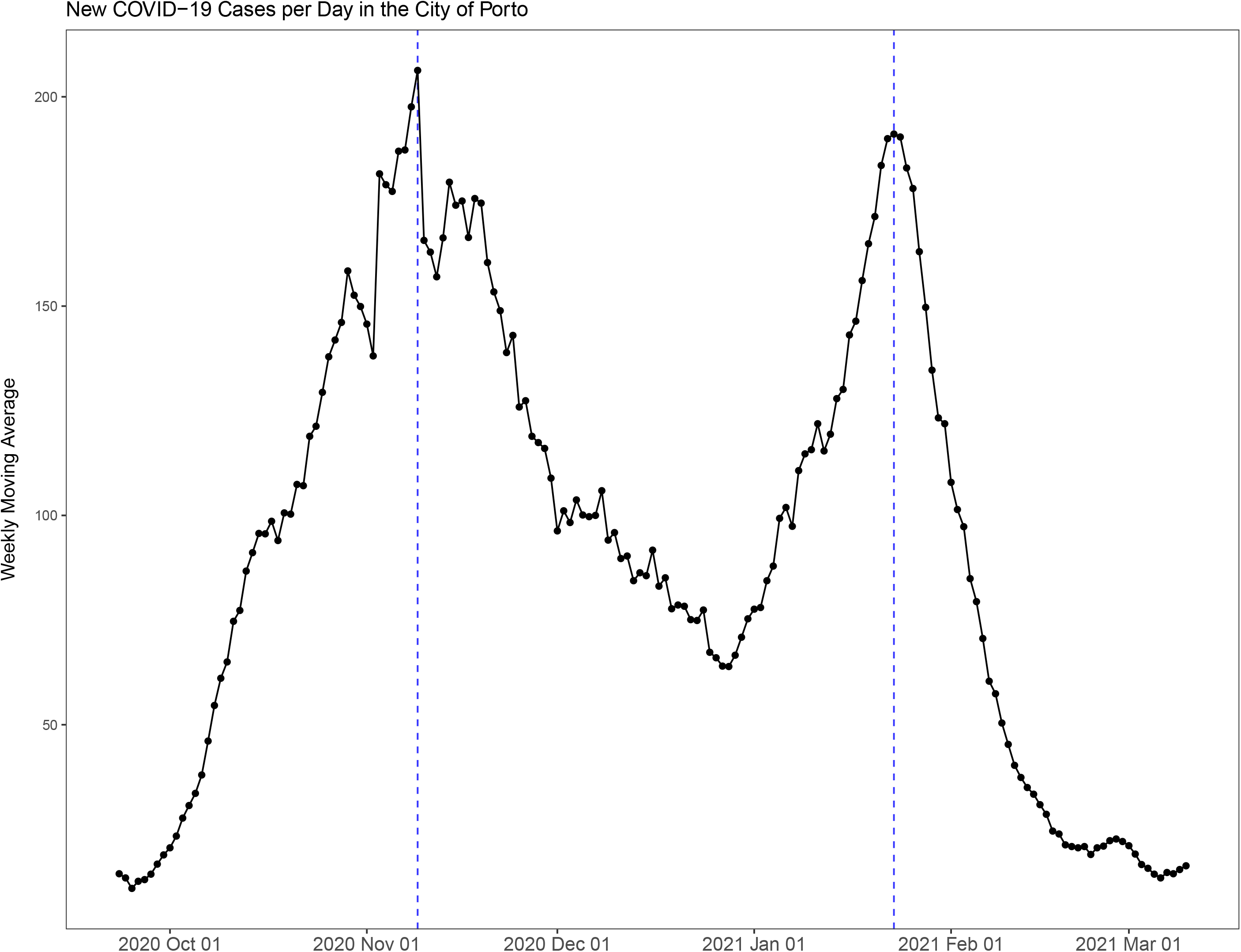
Seven-day moving average of new COVID-19 cases per day in the city of Porto during the second stage of this study (wastewater monitoring). Data source: Direção Geral de Saúde.

**Figure 5.**
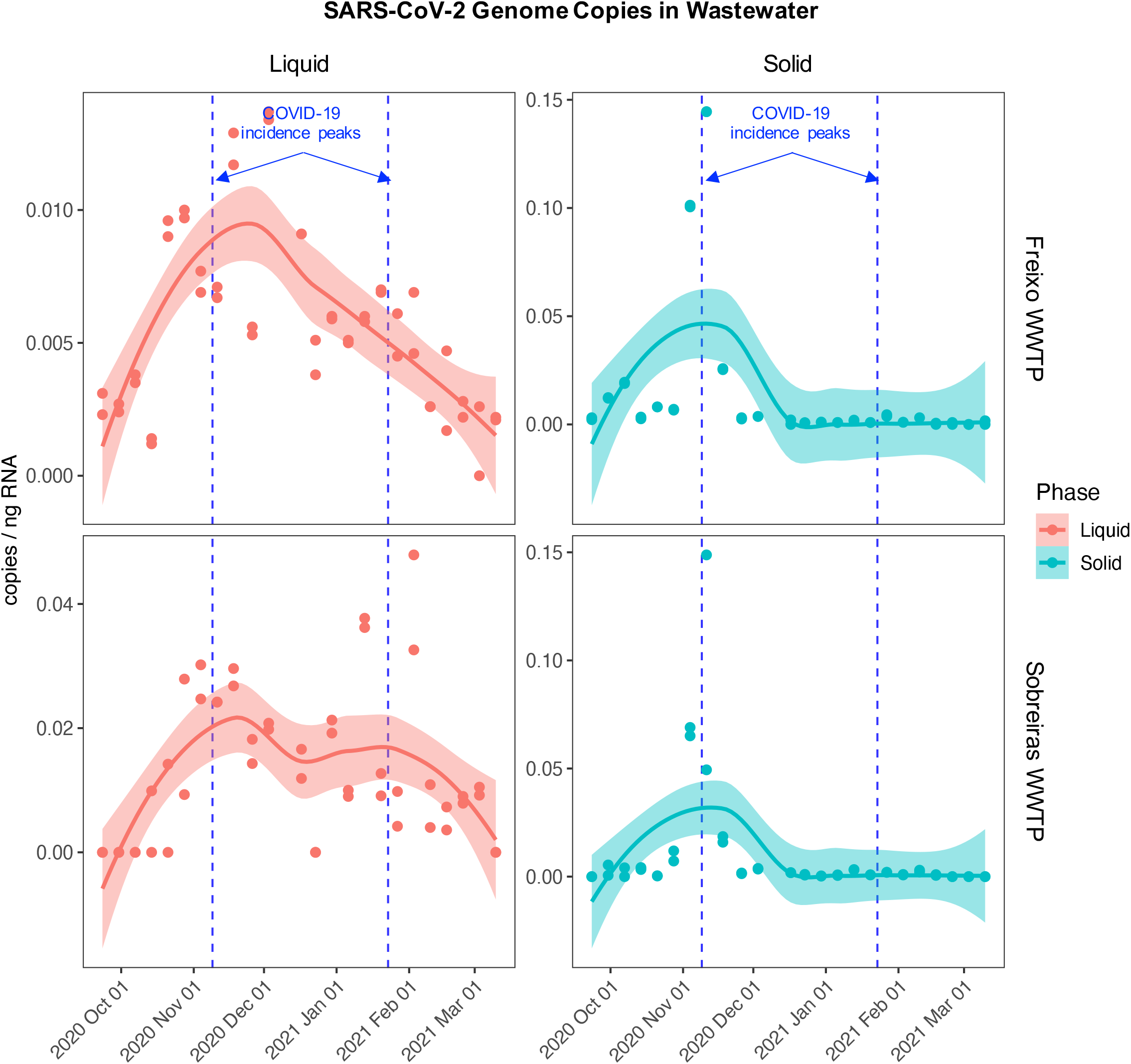
SARS-CoV-2 RNA abundance in the liquid and solid phases of untreated wastewater from Freixo and Sobreiras WWTPs in Porto, Portugal. The shaded area represents the 95% confidence interval of the locally estimated scatterplot smoothing predictions (solid line). The blue dashed lines represent the two COVID-19 incidence peaks observed in the city of Porto in mid-November and mid-January.

To further evaluate the apparent fit between SARS-CoV-2 RNA concentrations in wastewater and the COVID-19 cases in the city of Porto, simple linear regressions were performed (**Figure 6**). We found significant positive relationships between the SARS-CoV-2 copy numbers and the weekly moving average of COVID-19 cases in both phases of both WWTPs. The stronger associations were found in the liquid phase, especially in the Freixo WWTP, where 42% of the variation in the data was explained by this relationship. Similar relationships have been observed in previous studies in different world regions during the current pandemic (Medema et al., 2020; Peccia et al., 2020). Altogether, the results from this study support the use of the selected sampling and analysis workflow (method D, Figure 1) as a tool for long term monitoring of SARS-CoV-2 in wastewater. As suggested by Gonzalez et al. (2020), this sensitive wastewater monitoring may be used as a pre-screening tool to better target clinical testing. Additionally, this approach may be particularly suitable to complement clinical testing in evaluating temporal and spatial trends as well as monitoring the efficiency of public preventive measures. However, further investigation is needed to understand the conditions in which this approach may be more suitable to guide and support public health policies. For instance, knowing the population actively discharging to each monitored WWTP may be critical to link wastewater viral loads with cases of disease. Our ongoing project of tracking SARS-CoV-2 RNA in Porto WWTPs, together with other studies across the globe, will certainly contribute to answer these questions in the near future.

**Figure 6.**
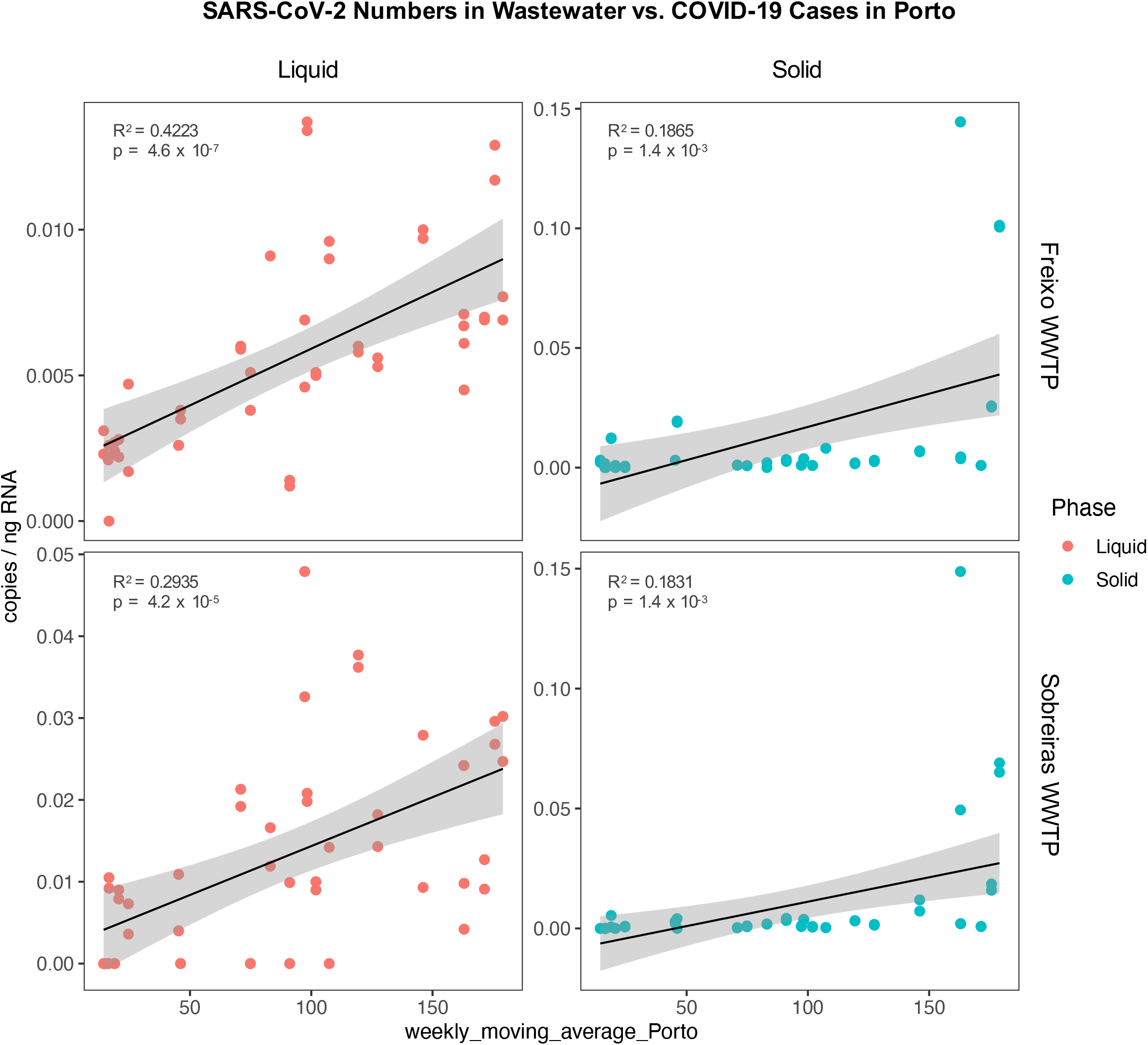
Linear regressions between SARS-CoV-2 RNA abundance in untreated wastewater and the 7-day moving average of new COVID-19 cases per day in the city of Porto. The shaded area represents the 95% confidence interval of the linear regression predictions.

## Data Availability

Detailed methods and data available upon request.

## Acknowledgments

This research was partially supported by national funds through FCT—Foundation for Science and Technology within the scope of UIDB/04423/2020 and UIDP/04423/2020. The authors are grateful for the invaluable laboratory assistance provided by Chiara Perrod, Catarina Fonseca, and Inês Nóbrega.

## Supplementary Material

**Table S1**. Detection of SARS-CoV-2 in wastewater samples during the second stage of this study (weekly monitoring). Table showing the qPCR Ct values, the total RNA concentrations (ng/μl), A260, A260/230, A260/280 in liquid and solid samples extracted by method D in Sobreiras and Freixo WWTPs (S_L= Sobreiras liquid; S_P= Sobreira solid; F_L= Freixo liquid; F_P= Freixo solid; A and B represent two instrument replicates from the same sample in qPCR analysis; ND = not detected).

**Figure S1**. Aerial photos of the sampling locations: A) Sobreiras WWTP, B) Freixo WWTP

