## Supplementary material for "Continuous monitoring of SARS-CoV-2 RNA in urban wastewater from Porto, Portugal: sampling and analysis protocols": Table S1

| Week | Date | WWTP | Phase | Sample ID | Ct | ng RNA/µL | A260 | A260/230 | A260/280 |
|---|---|---|---|---|---|---|---|---|---|
| 20 | 23/9/2020 | Freixo | Liquid | D20F_A | 33,60 | 122,50 | 3,06 | 0,59 | 1,63 |
| 20 | 23/9/2020 | Freixo | Liquid | D20F_B | 32,60 | - | - | - | - |
| 20 | 23/9/2020 | Sobreiras | Liquid | D20S_A | ND | 2,71 | 0,07 | 0,58 | 1,86 |
| 20 | 23/9/2020 | Sobreiras | Liquid | D20S_B | ND | - | - | - | - |
| 20 | 23/9/2020 | Freixo | Solid | D20F_P_A | 34,10 | 106,91 | 2,66 | 2,21 | 2,02 |
| 20 | 23/9/2020 | Freixo | Solid | D20F_P_B | 33,00 | - | - | - | - |
| 20 | 23/9/2020 | Sobreiras | Solid | D20S_P_A | ND | 27,80 | 0,70 | 1,25 | 1,98 |
| 20 | 23/9/2020 | Sobreiras | Solid | D20S_P_B | ND | - | - | - | - |
| 21 | 30/9/2020 | Freixo | Liquid | D21F_L_A | 34,10 | 90,54 | 2,26 | 0,43 | 1,44 |
| 21 | 30/9/2020 | Freixo | Liquid | D21F_L_B | 34,50 | - | - | - | - |
| 21 | 30/9/2020 | Sobreiras | Liquid | D21S_L_A | ND | 18,33 | 0,46 | 0,23 | 0,97 |
| 21 | 30/9/2020 | Sobreiras | Liquid | D21S_L_B | ND | - | - | - | - |
| 21 | 30/9/2020 | Freixo | Solid | D21F_P_A | 29,70 | 71,81 | 1,79 | 1,51 | 2,01 |
| 21 | 30/9/2020 | Freixo | Solid | D21F_P_B | 29,80 | - | - | - | - |
| 21 | 30/9/2020 | Sobreiras | Solid | D21S_P_A | 36,50 | 21,80 | 0,63 | 1,12 | 1,93 |
| 21 | 30/9/2020 | Sobreiras | Solid | D21S_P_B | 43,90 | - | - | - | - |
| 22 | 7/10/2020 | Freixo | Liquid | D22F_A | 31,20 | 165,30 | 4,13 | 0,54 | 1,48 |
| 22 | 7/10/2020 | Freixo | Liquid | D22F_B | 30,90 | - | - | - | - |
| 22 | 7/10/2020 | Sobreiras | Liquid | D22S_A | ND | 4,73 | 0,12 | 0,38 | 1,30 |
| 22 | 7/10/2020 | Sobreiras | Liquid | D22S_B | ND | - | - | - | - |
| 22 | 7/10/2020 | Freixo | Solid | D22F_P_A | 31,50 | 26,78 | 0,67 | 1,49 | 1,92 |
| 22 | 7/10/2020 | Freixo | Solid | D22F_P_B | 31,60 | - | - | - | - |
| 22 | 7/10/2020 | Sobreiras | Solid | D22S_P_A | ND | 13,47 | 0,34 | 0,66 | 2,53 |
| 22 | 7/10/2020 | Sobreiras | Solid | D22S_P_B | 39,00 | - | - | - | - |
| 23 | 14/10/2020 | Freixo | Liquid | D23F_A | 33,90 | 187,50 | 4,69 | 0,47 | 1,46 |
| 23 | 14/10/2020 | Freixo | Liquid | D23F_B | 34,20 | - | - | - | - |
| 23 | 14/10/2020 | Sobreiras | Liquid | D23S_A | 36,80 | 10,90 | 0,28 | 0,22 | 1,28 |
| 23 | 14/10/2020 | Sobreiras | Liquid | D23S_B | ND | - | - | - | - |
| 23 | 14/10/2020 | Freixo | Solid | D23F_P_A | 31,70 | 186,40 | 4,66 | 2,20 | 2,08 |
| 23 | 14/10/2020 | Freixo | Solid | D23F_P_B | 30,80 | - | - | - | - |
| 23 | 14/10/2020 | Sobreiras | Solid | D23S_P_A | 33,30 | 71,80 | 1,79 | 1,60 | 1,95 |
| 23 | 14/10/2020 | Sobreiras | Solid | D23S_P_B | 34,10 | - | - | - | - |
| 24 | 21/10/2020 | Freixo | Liquid | D24F_A | 29,60 | 102,10 | | | |
| 24 | 21/10/2020 | Freixo | Liquid | D24F_B | 29,40 | - | - | - | - |
| 24 | 21/10/2020 | Sobreiras | Liquid | D24S_A | 34,90 | 13,30 | | | |
| 24 | 21/10/2020 | Sobreiras | Liquid | D24S_B | ND | - | - | - | - |
| 24 | 21/10/2020 | Freixo | Solid | D24F_P_A | 29,40 | 121,00 | | | |
| 24 | 21/10/2020 | Freixo | Solid | D24F_P_B | 29,40 | - | - | - | - |
| 24 | 21/10/2020 | Sobreiras | Solid | D24S_P_A | 40,90 | 94,90 | | | |
| 24 | 21/10/2020 | Sobreiras | Solid | D24S_P_B | 40,10 | - | - | - | - |
| 25 | 28/10/2020 | Freixo | Liquid | D25F_A | 29,70 | 89,50 | 2,38 | 1,49 | |
| 25 | 28/10/2020 | Freixo | Liquid | D25F_B | 29,80 | - | - | - | - |
| 25 | 28/10/2020 | Sobreiras | Liquid | D25S_A | 33,50 | 10,30 | 0,26 | 1,49 | 0,51 |
| 25 | 28/10/2020 | Sobreiras | Liquid | D25S_B | 37,20 | - | - | - | - |
| 25 | 28/10/2020 | Freixo | Solid | D25F_P_A | 29,70 | 135,40 | 3,38 | 2,07 | 7,35 |
| 25 | 28/10/2020 | Freixo | Solid | D25F_P_B | 29,50 | - | - | - | - |
| 25 | 28/10/2020 | Sobreiras | Solid | D25S_P_A | 34,60 | 28,80 | 0,72 | 1,92 | 1,91 |
| 25 | 28/10/2020 | Sobreiras | Solid | D25S_P_B | 32,90 | - | - | - | - |
| 26 | 4/11/2020 | Freixo | Liquid | D26F_A | 29,51 | 121,70 | 3,04 | 0,53 | 1,49 |
| 26 | 4/11/2020 | Freixo | Liquid | D26F_B | 29,90 | - | - | - | - |
| 26 | 4/11/2020 | Sobreiras | Liquid | D26SS_A | 31,43 | 17,64 | 0,44 | 0,53 | 1,34 |
| 26 | 4/11/2020 | Sobreiras | Liquid | D26SS_B | 32,10 | - | - | - | - |
| 26 | 4/11/2020 | Freixo | Solid | D26F_P_A | 28,76 | 11,66 | 0,29 | 1,19 | 1,72 |
| 26 | 4/11/2020 | Freixo | Solid | D26F_P_B | 28,79 | - | - | - | - |
| 26 | 4/11/2020 | Sobreiras | Solid | D26S_P_A | 30,76 | 10,00 | 0,25 | 1,15 | 1,52 |
| 26 | 4/11/2020 | Sobreiras | Solid | D26S_P_B | 30,56 | - | - | - | - |

| Week | Date | WWTP | Phase | Sample ID | Ct | ng RNA/µL | A260 | A260/230 | A260/280 |
|---|---|---|---|---|---|---|---|---|---|
| 27 | 11/11/2020 | Freixo | Liquid | D27F_A | 31,30 | 82,74 | 2,07 | 0,59 | 1,46 |
| 27 | 11/11/2020 | Freixo | Liquid | D27F_B | 31,10 | - | - | - | - |
| 27 | 11/11/2020 | Sobreiras | Liquid | D27SS_A | 33,80 | 10,84 | 0,27 | 0,41 | 1,43 |
| 27 | 11/11/2020 | Sobreiras | Liquid | D27SS_B | 33,80 | - | - | - | - |
| 27 | 11/11/2020 | Freixo | Solid | D27F_P_A | 30,70 | 4,58 | 0,11 | 0,75 | 1,47 |
| 27 | 11/11/2020 | Freixo | Solid | D27F_P_B | 30,70 | - | - | - | - |
| 27 | 11/11/2020 | Sobreiras | Solid | D27S_P_A | 33,30 | 2,05 | 0,05 | 0,30 | 0,99 |
| 27 | 11/11/2020 | Sobreiras | Solid | D27S_P_B | 37,00 | - | - | - | - |
| 28 | 18/11/2020 | Freixo | Liquid | D28F_A | 31,18 | 48,85 | 1,22 | 0,55 | 1,43 |
| 28 | 18/11/2020 | Freixo | Liquid | D28F_B | 30,87 | - | - | - | - |
| 28 | 18/11/2020 | Sobreiras | Liquid | D28SS_A | 32,31 | 15,29 | 0,38 | 1,11 | 1,54 |
| 28 | 18/11/2020 | Sobreiras | Liquid | D28SS_B | 31,98 | - | - | - | - |
| 28 | 18/11/2020 | Freixo | Solid | D28F_P_A | 30,67 | 25,85 | 0,65 | 1,36 | 1,77 |
| 28 | 18/11/2020 | Freixo | Solid | D28F_P_B | 30,75 | - | - | - | - |
| 28 | 18/11/2020 | Sobreiras | Solid | D28S_P_A | 29,91 | 45,20 | 1,13 | 1,63 | 1,83 |
| 28 | 18/11/2020 | Sobreiras | Solid | D28S_P_B | 30,42 | - | - | - | - |
| 29 | 26/11/2020 | Freixo | Liquid | D29F_L_A | 30,94 | 115,70 | 2,89 | 0,61 | 1,61 |
| 29 | 26/11/2020 | Freixo | Liquid | D29F_L_B | 30,77 | - | - | - | - |
| 29 | 26/11/2020 | Sobreiras | Liquid | D29S_L_A | 33,60 | 19,80 | 0,49 | 0,86 | 1,50 |
| 29 | 26/11/2020 | Sobreiras | Liquid | D29S_L_B | 32,74 | - | - | - | - |
| 29 | 26/11/2020 | Freixo | Solid | D29F_P_A | 32,01 | 179,60 | 4,49 | 2,13 | 2,04 |
| 29 | 26/11/2020 | Freixo | Solid | D29F_P_B | 31,32 | - | - | - | - |
| 29 | 26/11/2020 | Sobreiras | Solid | D29S_P_A | 35,54 | 93,30 | 2,54 | 1,84 | 2,02 |
| 29 | 26/11/2020 | Sobreiras | Solid | D29S_P_B | 36,22 | - | - | - | - |
| 30 | 3/12/2020 | Freixo | Liquid | D30F_L_A | 28,40 | 98,40 | 2,46 | 0,64 | 1,54 |
| 30 | 3/12/2020 | Freixo | Liquid | D30F_L_B | 28,30 | - | - | - | - |
| 30 | 3/12/2020 | Sobreiras | Liquid | D30S_L_A | 33,30 | 15,60 | 0,39 | 0,80 | 1,56 |
| 30 | 3/12/2020 | Sobreiras | Liquid | D30S_L_B | 33,10 | - | - | - | - |
| 30 | 3/12/2020 | Freixo | Solid | D30F_P_A | 29,00 | 294,50 | 7,36 | 2,39 | 2,08 |
| 30 | 3/12/2020 | Freixo | Solid | D30F_P_B | 29,10 | - | - | - | - |
| 30 | 3/12/2020 | Sobreiras | Solid | D30S_P_A | 29,60 | 261,90 | 6,55 | 2,34 | 2,07 |
| 30 | 3/12/2020 | Sobreiras | Solid | D30S_P_B | 29,30 | - | - | - | - |
| 32 | 17/12/2020 | Freixo | Liquid | D32F_L_A | error | 66,20 | 1,66 | 0,69 | 1,53 |
| 32 | 17/12/2020 | Freixo | Liquid | D32F_L_B | 31,00 | - | - | - | - |
| 32 | 17/12/2020 | Sobreiras | Liquid | D32S_L_A | 32,00 | 27,10 | 0,68 | 0,65 | 1,41 |
| 32 | 17/12/2020 | Sobreiras | Liquid | D32S_L_B | 33,10 | - | - | - | - |
| 32 | 17/12/2020 | Freixo | Solid | D32F_P_A | 30,00 | 379,50 | 9,49 | 2,36 | 2,11 |
| 32 | 17/12/2020 | Freixo | Solid | D32F_P_B | 30,20 | - | - | - | - |
| 32 | 17/12/2020 | Sobreiras | Solid | D32S_P_A | 30,20 | 417,40 | 10,44 | 2,49 | 2,11 |
| 32 | 17/12/2020 | Sobreiras | Solid | D32S_P_B | 30,20 | - | - | - | - |
| 33 | 23/12/2020 | Freixo | Liquid | D33F_L_A | 35,00 | 49,10 | 1,20 | 0,30 | 1,30 |
| 33 | 23/12/2020 | Freixo | Liquid | D33F_L_B | 33,90 | - | - | - | - |
| 33 | 23/12/2020 | Sobreiras | Liquid | D33S_L_A | ND | 9,90 | 0,24 | 0,55 | 1,50 |
| 33 | 23/12/2020 | Sobreiras | Liquid | D33S_L_B | ND | - | - | - | - |
| 33 | 23/12/2020 | Freixo | Solid | D33F_P_A | 32,90 | 414,50 | 10,36 | 2,20 | 2,09 |
| 33 | 23/12/2020 | Freixo | Solid | D33F_P_B | 32,80 | - | - | - | - |
| 33 | 23/12/2020 | Sobreiras | Solid | D33S_P_A | 34,60 | 196,60 | 4,90 | 2,16 | 2,03 |
| 33 | 23/12/2020 | Sobreiras | Solid | D33S_P_B | 35,90 | - | - | - | - |
| 34 | 30/12/2020 | Freixo | Liquid | D34F_L_A | 31,50 | 86,50 | 2,16 | 0,65 | 1,61 |
| 34 | 30/12/2020 | Freixo | Liquid | D34F_L_B | 31,50 | - | - | - | - |
| 34 | 30/12/2020 | Sobreiras | Liquid | D34S_L_A | 33,20 | 16,40 | 0,41 | 0,89 | 1,58 |
| 34 | 30/12/2020 | Sobreiras | Liquid | D34S_L_B | 32,80 | - | - | - | - |
| 34 | 30/12/2020 | Freixo | Solid | D34F_P_A | 32,00 | 458,10 | 11,45 | 2,46 | 2,10 |
| 34 | 30/12/2020 | Freixo | Solid | D34F_P_B | 31,50 | - | - | - | - |
| 34 | 30/12/2020 | Sobreiras | Solid | D34S_P_A | 32,50 | 1104,90 | 27,62 | 2,54 | 2,16 |
| 34 | 30/12/2020 | Sobreiras | Solid | D34S_P_B | 33,90 | - | - | - | - |
| 35 | 6/1/2021 | Freixo | Liquid | D35F_L_A | 31,50 | 102,20 | | | |

| Week | Date | WWTP | Phase | Sample ID | Ct | ng RNA/µL | A260 | A260/230 | A260/280 |
|---|---|---|---|---|---|---|---|---|---|
| 35 | 6/1/2021 | Freixo | Liquid | D35F_L_B | 31,60 | - | - | - | - |
| 35 | 6/1/2021 | Sobreiras | Liquid | D35S_L_A | 33,70 | 30,00 | | | |
| 35 | 6/1/2021 | Sobreiras | Liquid | D35S_L_B | 33,40 | - | - | - | - |
| 35 | 6/1/2021 | Freixo | Solid | D35F_P_A | 31,50 | 558,40 | | | |
| 35 | 6/1/2021 | Freixo | Solid | D35F_P_B | 31,50 | - | - | - | - |
| 35 | 6/1/2021 | Sobreiras | Solid | D35S_P_A | 34,00 | 469,80 | | | |
| 35 | 6/1/2021 | Sobreiras | Solid | D35S_P_B | 32,50 | - | - | - | - |
| 36 | 13/1/2021 | Freixo | Liquid | D36F_L_A | 32,30 | 71,20 | 1,78 | 0,65 | 1,53 |
| 36 | 13/1/2021 | Freixo | Liquid | D36F_L_B | 32,10 | - | - | - | - |
| 36 | 13/1/2021 | Sobreiras | Liquid | D36S_L_A | 32,80 | 9,70 | 0,24 | 0,97 | 1,67 |
| 36 | 13/1/2021 | Sobreiras | Liquid | D36S_L_B | 32,70 | - | - | - | - |
| 36 | 13/1/2021 | Freixo | Solid | D36F_P_A | 32,00 | 222,90 | 5,57 | 2,16 | 2,10 |
| 36 | 13/1/2021 | Freixo | Solid | D36F_P_B | 32,80 | - | - | - | - |
| 36 | 13/1/2021 | Sobreiras | Solid | D36S_P_A | 32,10 | 138,70 | 3,47 | 2,28 | 2,05 |
| 36 | 13/1/2021 | Sobreiras | Solid | D36S_P_B | 32,00 | - | - | - | - |
| 37 | 20/1/2021 | Freixo | Liquid | D37F_L_A | 30,80 | 93,20 | 2,30 | 0,69 | 1,64 |
| 37 | 20/1/2021 | Freixo | Liquid | D37F_L_B | 30,80 | - | - | - | - |
| 37 | 20/1/2021 | Sobreiras | Liquid | D37S_L_A | 33,30 | 23,90 | 0,50 | 0,72 | 1,33 |
| 37 | 20/1/2021 | Sobreiras | Liquid | D37S_L_B | 34,40 | - | - | - | - |
| 37 | 20/1/2021 | Freixo | Solid | D37F_P_A | 32,20 | 495,20 | 12,38 | 2,49 | 2,14 |
| 37 | 20/1/2021 | Freixo | Solid | D37F_P_B | 32,00 | - | - | - | - |
| 37 | 20/1/2021 | Sobreiras | Solid | D37S_P_A | 32,30 | 466,90 | 11,65 | 2,48 | 2,17 |
| 37 | 20/1/2021 | Sobreiras | Solid | D37S_P_B | 32,70 | - | - | - | - |
| 38 | 27/1/2021 | Freixo | Liquid | D38F_L_A | 32,00 | 98,60 | 2,46 | 0,73 | 1,64 |
| 38 | 27/1/2021 | Freixo | Liquid | D38F_L_B | 31,00 | - | - | - | - |
| 38 | 27/1/2021 | Sobreiras | Liquid | D38S_L_A | 37,00 | 24,50 | 0,61 | 0,73 | 1,51 |
| 38 | 27/1/2021 | Sobreiras | Liquid | D38S_L_B | 34,10 | - | - | - | - |
| 38 | 27/1/2021 | Freixo | Solid | D38F_P_A | 31,90 | 125,00 | 3,15 | 2,18 | 1,99 |
| 38 | 27/1/2021 | Freixo | Solid | D38F_P_B | 31,30 | - | - | - | - |
| 38 | 27/1/2021 | Sobreiras | Solid | D38S_P_A | 31,80 | 238,10 | 5,95 | 2,35 | 2,06 |
| 38 | 27/1/2021 | Sobreiras | Solid | D38S_P_B | 31,80 | - | - | - | - |
| 39 | 3/2/2021 | Freixo | Liquid | D39F_L_A | 32,54 | 55,43 | 1,38 | 0,88 | 1,51 |
| 39 | 3/2/2021 | Freixo | Liquid | D39F_L_B | 33,90 | - | - | - | - |
| 39 | 3/2/2021 | Sobreiras | Liquid | D39S_L_A | 33,67 | 8,39 | 0,21 | 0,87 | 1,41 |
| 39 | 3/2/2021 | Sobreiras | Liquid | D39S_L_B | 32,38 | - | - | - | - |
| 39 | 3/2/2021 | Freixo | Solid | D39F_P_A | 32,09 | 447,28 | 11,18 | 2,52 | 2,18 |
| 39 | 3/2/2021 | Freixo | Solid | D39F_P_B | 32,11 | - | - | - | - |
| 39 | 3/2/2021 | Sobreiras | Solid | D39S_P_A | 31,03 | 591,31 | 14,78 | 2,53 | 2,13 |
| 39 | 3/2/2021 | Sobreiras | Solid | D39S_P_B | 31,97 | - | - | - | - |
| 40 | 10/2/2021 | Freixo | Liquid | D40F_L_A | 34,39 | 84,25 | 2,11 | 0,54 | 1,41 |
| 40 | 10/2/2021 | Freixo | Liquid | D40F_L_B | 34,44 | - | - | - | - |
| 40 | 10/2/2021 | Sobreiras | Liquid | D40S_L_A | 34,38 | 20,34 | 0,51 | 0,68 | 1,43 |
| 40 | 10/2/2021 | Sobreiras | Liquid | D40S_L_B | 37,72 | - | - | - | - |
| 40 | 10/2/2021 | Freixo | Solid | D40F_P_A | 35,43 | 52,94 | 1,32 | 1,76 | 1,91 |
| 40 | 10/2/2021 | Freixo | Solid | D40F_P_B | error | - | - | - | - |
| 40 | 10/2/2021 | Sobreiras | Solid | D40S_P_A | 35,01 | 85,99 | 2,15 | 1,95 | 1,96 |
| 40 | 10/2/2021 | Sobreiras | Solid | D40S_P_B | 33,89 | - | - | - | - |
| 41 | 17/2/2021 | Freixo | Liquid | D41F_L_A | 37,97 | 44,98 | 1,12 | 0,80 | 1,58 |
| 41 | 17/2/2021 | Freixo | Liquid | D41F_L_B | 34,54 | - | - | - | - |
| 41 | 17/2/2021 | Sobreiras | Liquid | D41S_L_A | 37,59 | 23,90 | 0,59 | 1,96 | 1,64 |
| 41 | 17/2/2021 | Sobreiras | Liquid | D41S_L_B | 35,17 | - | - | - | - |
| 41 | 17/2/2021 | Freixo | Solid | D41F_P_A | ND | 137,50 | 3,43 | 1,72 | 2,09 |
| 41 | 17/2/2021 | Freixo | Solid | D41F_P_B | 37,45 | - | - | - | - |
| 41 | 17/2/2021 | Sobreiras | Solid | D41S_P_A | 36,61 | 197,30 | 4,49 | 1,64 | 1,96 |
| 41 | 17/2/2021 | Sobreiras | Solid | D41S_P_B | 35,01 | - | - | - | - |
| 42 | 24/2/2021 | Freixo | Liquid | D42F_L_A | 33,37 | 105,86 | 2,65 | 0,62 | 1,54 |
| 42 | 24/2/2021 | Freixo | Liquid | D42F_L_B | 34,14 | - | - | - | - |

| Week | Date | WWTP | Phase | Sample ID | Ct | ng RNA/µL | A260 | A260/230 | A260/280 |
|---|---|---|---|---|---|---|---|---|---|
| 42 | 24/2/2021 | Sobreiras | Liquid | D42S_L_A | 34,91 | 20,96 | 0,52 | 0,71 | 1,48 |
| 42 | 24/2/2021 | Sobreiras | Liquid | D42S_L_B | 35,33 | - | - | - | - |
| 42 | 24/2/2021 | Freixo | Solid | D42F_P_A | 40,99 | 35,02 | 0,88 | 1,76 | 1,93 |
| 42 | 24/2/2021 | Freixo | Solid | D42F_P_B | ND | - | - | - | - |
| 42 | 24/2/2021 | Sobreiras | Solid | D42S_P_A | ND | 91,70 | 2,29 | 2,28 | 2,00 |
| 42 | 24/2/2021 | Sobreiras | Solid | D42S_P_B | ND | - | - | - | - |
| 43 | 3/3/2021 | Freixo | Liquid | D43F_L_A | ND | 106,45 | 2,66 | 0,60 | 1,52 |
| 43 | 3/3/2021 | Freixo | Liquid | D43F_L_B | 33,60 | - | - | - | - |
| 43 | 3/3/2021 | Sobreiras | Liquid | D43S_L_A | 35,33 | 18,04 | 0,45 | 2,40 | 1,70 |
| 43 | 3/3/2021 | Sobreiras | Liquid | D43S_L_B | 34,89 | - | - | - | - |
| 43 | 3/3/2021 | Freixo | Solid | D43F_P_A | ND | 102,66 | 2,56 | 2,63 | 2,01 |
| 43 | 3/3/2021 | Freixo | Solid | D43F_P_B | ND | - | - | - | - |
| 43 | 3/3/2021 | Sobreiras | Solid | D43S_P_A | ND | 63,63 | 1,59 | 2,62 | 2,07 |
| 43 | 3/3/2021 | Sobreiras | Solid | D43S_P_B | ND | - | - | - | - |
| 44 | 10/3/2021 | Freixo | Liquid | D44F_L_A | 34,64 | 93,78 | 2,34 | 0,60 | 1,56 |
| 44 | 10/3/2021 | Freixo | Liquid | D44F_L_B | 34,81 | - | - | - | - |
| 44 | 10/3/2021 | Sobreiras | Liquid | D44S_L_A | ND | 13,15 | 0,33 | 0,44 | 1,50 |
| 44 | 10/3/2021 | Sobreiras | Liquid | D44S_L_B | ND | - | - | - | - |
| 44 | 10/3/2021 | Freixo | Solid | D44F_P_A | 36,99 | 59,66 | 1,49 | 2,05 | 1,95 |
| 44 | 10/3/2021 | Freixo | Solid | D44F_P_B | ND | - | - | - | - |
| 44 | 10/3/2021 | Sobreiras | Solid | D44S_P_A | ND | 61,43 | 1,54 | 2,03 | 1,96 |
