## Supplementary figures and images for "Continuous monitoring of SARS-CoV-2 RNA in urban wastewater from Porto, Portugal: sampling and analysis protocols"

### Figure S1

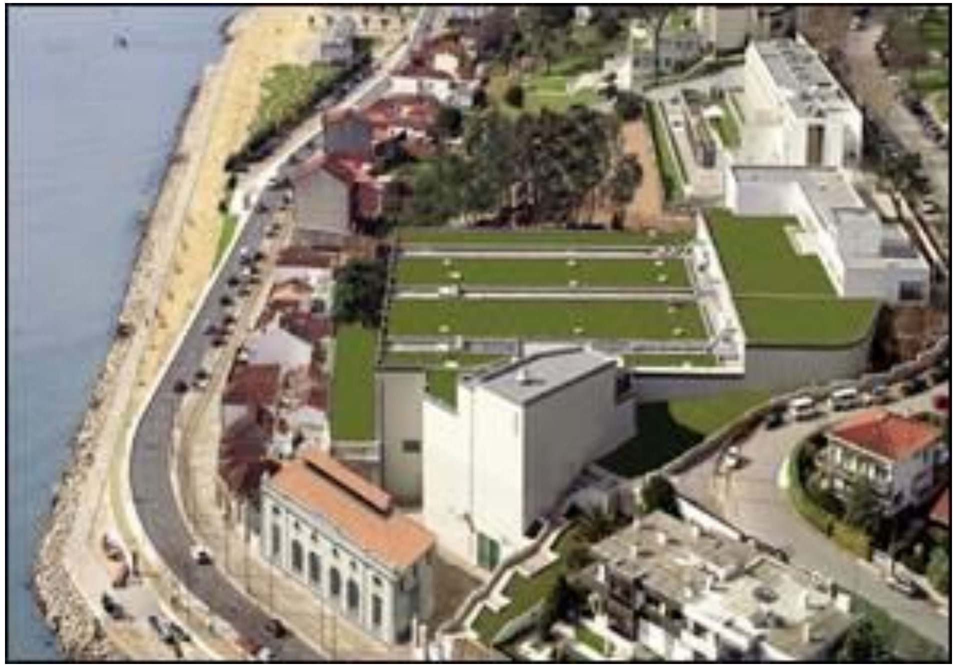

A

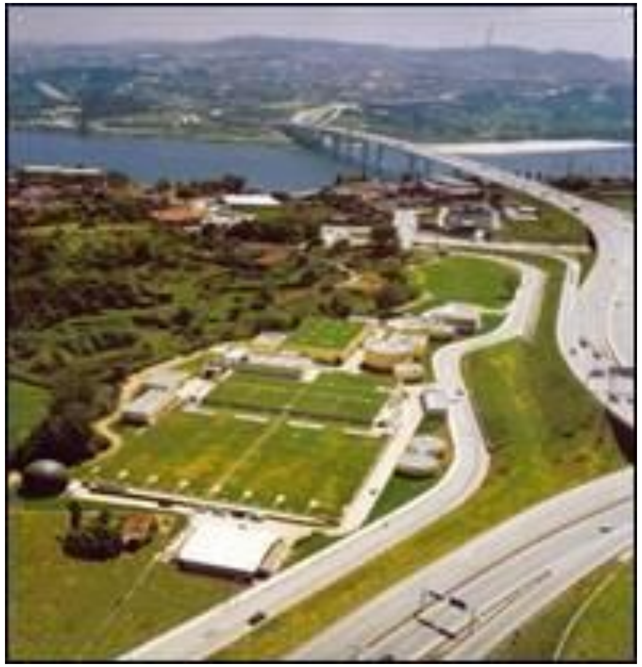

B
